# COVID19-Global: A shiny application to perform a global comparative data visualization for the SARS-CoV-2 epidemic

**DOI:** 10.1101/2020.05.18.20105684

**Authors:** Aurelio Tobías, Pau Satorra, Joan Valls, Cristian Tebé

## Abstract

Data visualization is an essential tool for exploring and communicating findings in medical research, especially in epidemiological surveillance. The COVID19-Global online web application systematically produces daily updated data visualization and analysis of the SARS-CoV-2 epidemic on a global scale. It collects automatically daily data on COVID-19 diagnosed cases and mortality worldwide from January 1^st^, 2020 onwards. We have implemented comparative data visualization between countries for the most common indicators in epidemiological surveillance to follow an epidemic: attack rate, population fatality rate, case fatality rate, and basic reproduction number. The application may help for a better understanding of the SARS-CoV-2 epidemic worldwide.

## 1. INTRODUCTION

The first confirmed case of SARS-CoV-2 in China was reported to the WHO country office in China on December 31^st^, 2019 (1). The outbreak was declared a public health emergency of international concern on January 30^th^, 2020 (1). Since then, 215 countries have been affected worldwide, 4,722,233 people have been diagnosed cases, and 313,266 have died due to the SARS-CoV-2 pandemic (2).

Data visualization and analysis is an essential tool for exploring and communicating findings in medical research, and especially in epidemiological surveillance (3). It can help researchers and policymakers identify trends that could be overlooked if the data were reviewed in tabular form. We have developed a Shiny application to compare epidemiological indicators on the SARS-CoV-2 epidemic.

## 2. SOFTWARE AVAILABILITY AND REQUIREMENTS

The COVID19-Tracker app has been developed in RStudio (4), version 1.2.5033, using the Shiny package, version 1.4.0. Shiny offers the ability to develop a graphical user interface (GUI) that can be run locally or deployed online. Last is particularly beneficial to show and communicate updated findings to a broad audience. All the analyses have been carried out using R (5), version 3.6.3.

The application has a friendly structure based on menus to shown data visualization for the most common indicators in epidemiological surveillance to follow an epidemic: attack rate, population fatality rate, case fatality rate, and basic reproduction number (Figure 1).

**Figure 1.**
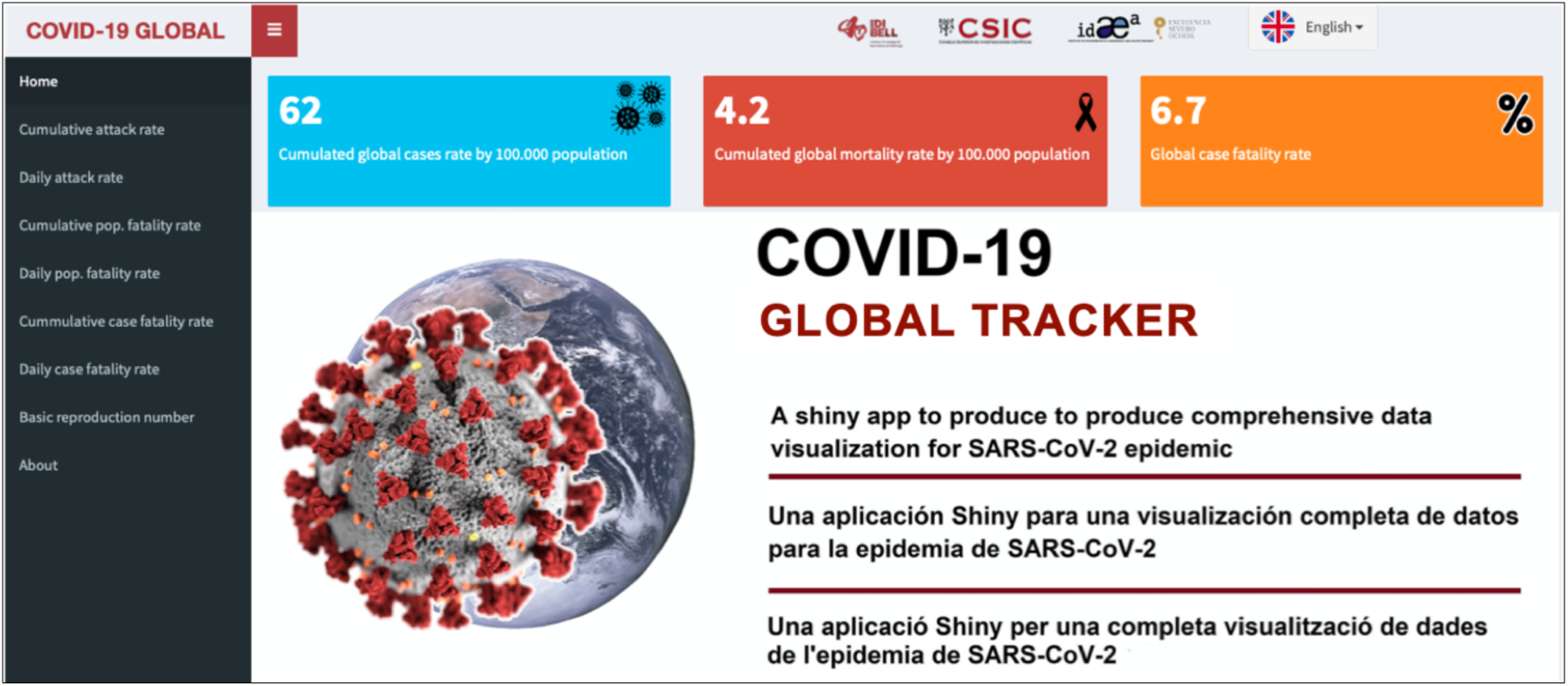
Home page of the COVID19-Global application, for visualization and analysis of data from the SARS-CoV-2 worldwied. Available at: https://ubidi.shinvapps.io/covidl9global/

Two additional menus are already implemented to describe the epidemiological indicators analyses and collecting other applications, also developed in Shiny, by other users to follow the COVID19 epidemic globally.

The app has an automated process to update data and all analyses every time a user connects to the app. It is available online at the following link: https://ubidi.shinvapps.io/covidl9global/ and shortly free available on Github as an R package. The application allows comparing epidemiological indicators between countries on the current date or since the start of the epidemic in each country. The displayed graphs are mouse-sensitive, showing the observed and expected number of events through the plot. The graphs can also be displayed on a log scale. Likewise, when selecting any plot, the application allows the option of downloading it as a portable network graphic (*.png). All menus are available in English, Spanish, and Catalan.

## 3. DATA SOURCES

We collected daily data on COVID-19 diagnosed cases and mortality, from January I^s^, 2020, onwards. Data is collected automatically from the ECDC’s (European Centre for Disease Prevention and Control) the geographical distribution of COVID-19 cases worldwide (6). The downloadable dataset is updated daily and contains the latest available public data on COVID-19 worldwide.

## 4. METHODS

We have implemented a data visualization for the following epidemiological indicators:

The *attack rate* is the ratio between the positively diagnosed cases (T+) and the total population (P) in a given country (AR = C+/P).

The *population fatality rate* is the ratio between the positively diagnosed deaths (D+) and the population (P) in a given country (PFR = D+/P).

The *case fatality rate* is the ratio between the positively diagnosed deaths (D+) and the positively tested cases (C+) in a given country (CFR = D+/C+).

The *basic reproduction number* (R_0_) is the average number of secondary cases of disease caused by a single infected individual over his or her infectious period (7). Here, we used the R package *EpiEstim* to estimate the R_0_ (7).

Figure 2 shows an example of these indicators comparing the six most affected countries worldwide (United States, Russia, Brazil, Spain, United Kingdom and Italy up to May 17^th^, 2020) since the epidemic started in each country.

**Figure 2.**
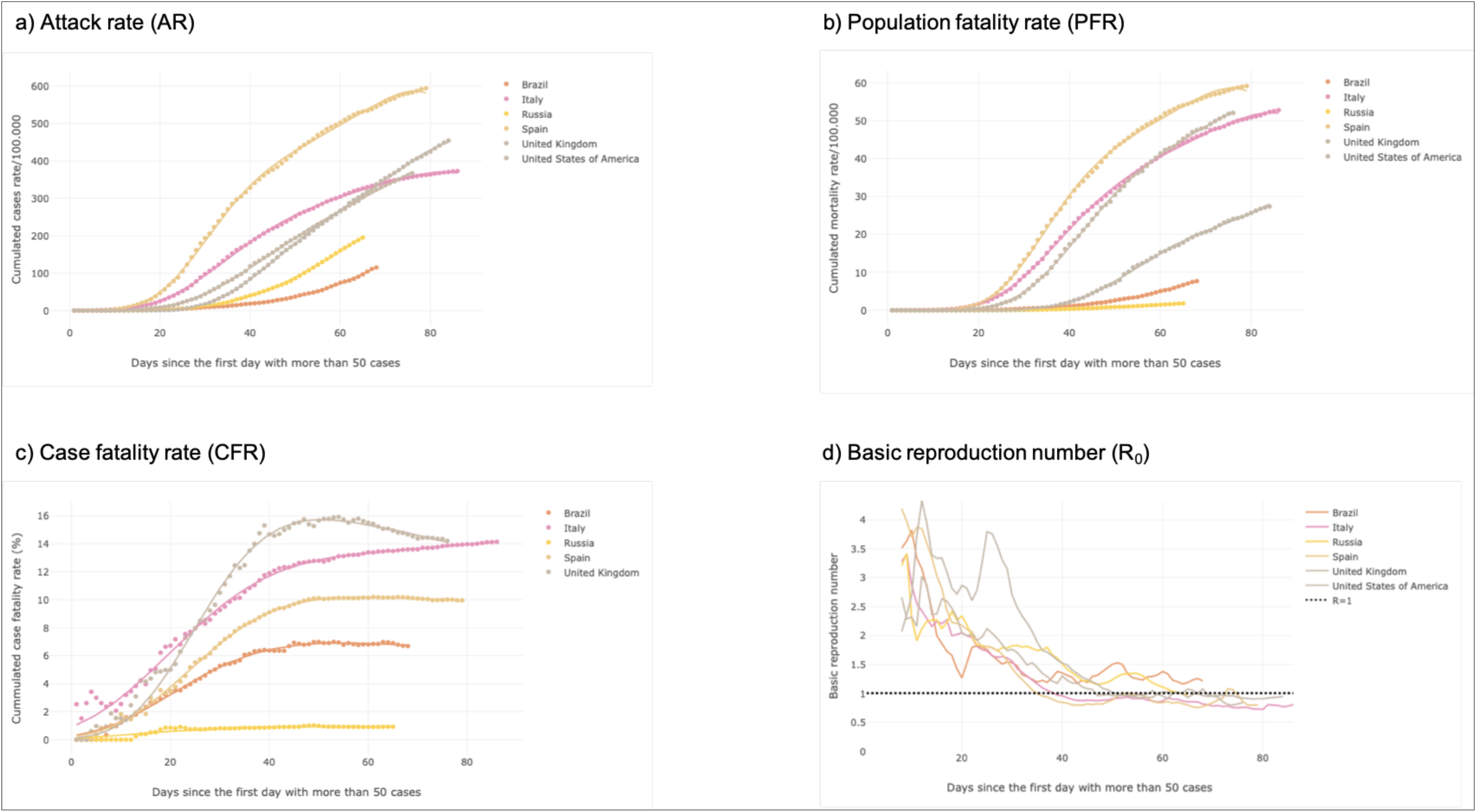
Standard output display of the COVID19-Global application (results updated to May 17^th^, 2020) for the attack rate (a), population fatality rate (b), case fatality rate (c), and basic reproduction number (R_0_), for the six countries wit the largest number of diagnosed cases (United States, Russia, Brazil, Spain, United Kindom and Italy).

## 5. DISCUSSION

Disease surveillance forms the basis for response COVID-19 epidemic and requires knowing trends in disease frequency in different subgroups and locations. The COVID19-Global application provides a global overview for the epidemiological surveillance of the pandemic worldwide, visualizing in a simple and intuitive way the main epidemiological indicators of all countries affected by the SARS-CoV-2 pandemic with daily updated data.

Country comparisons based on counts and their trends across populations and places should be replaced by rate comparisons adjusting the count to the size of the population (8). However, we should acknowledge that it is not possible to make an accurate estimate of the rates due to the underreporting of diagnosed cases and mortality in official statistics (9). Moreover, the application does not take into account the changes in the definition of diagnosed cases, nor the lockdown measures are undertaken in each country, aiming to flatten the curve. Moreover, the selection of the number of people who have been tested is critical for an accurate estimation (8). Accurate estimation of the rates depends on the testing strategy, the prevalence of infection, and the test accuracy. Differences between countries or overtime may merely reflect differences in selection for testing and test performance (8). In any case, a routine health system data of basic epidemiological indicators for the SARS-CoV-2 pandemic allowing for the comparison between countries, is essential for surveillance epidemiology and health policy.

We continue to plan improvements to the application to include specific data visualizations by country and aggregated by geographical regions. In summary, this application, easy to use, comes to fill a gap in this particular scenario for the visualization of epidemiological data for the COVID-19 at a global scale.

## Data Availability

Daily data on COVID-19 diagnosed cases and mortality, from January 1s, 2020, onwards, is collected automatically from the ECDC’s (European Centre for Disease Prevention and Control) the geographical distribution of COVID-19 cases worldwide.

https://www.ecdc.europa.eu/en/publications-data/download-todays-data-geographic-distribution-covid-19-cases-worldwide/

## Acknowledgements

None.

